# Effectiveness of Influenza Vaccine for Preventing Laboratory-Confirmed Influenza Hospitalizations in Immunocompromised Adults

**DOI:** 10.1101/2020.10.08.20208579

**Authors:** Kailey Hughes, Donald B Middleton, Mary Patricia Nowalk, Goundappa K Balasubramani, Emily T Martin, Manjusha Gaglani, H Keipp Talbot, Manish M Patel, Jill M Ferdinands, Richard K Zimmerman, Fernanda P Silveira, for the HAIVEN Study Investigators

**Author notes:** **Corresponding author:** Fernanda P. Silveira, MD, MS, FIDSA, Associate Professor of Medicine, 3601 Fifth Avenue, Suite 5B, Pittsburgh, PA 15213, **Alternate corresponding author:** Kailey Hughes, MPH, 3601 Fifth Avenue, Suite 5B, Pittsburgh, PA 15213.

## Abstract

**Background:** Yearly influenza immunization is recommended for immunocompromised (IC) individuals, although immune responses are lower than that for the non-immunocompromised and the data on vaccine effectiveness (VE) in the IC is scarce. We evaluated VE against influenza-associated hospitalization among IC adults.

**Methods:** We analyzed data from adults ≥ 18 years hospitalized with acute respiratory illness (ARI) during the 2017-2018 influenza season at 10 hospitals in the United States. IC adults were identified using pre-specified case-definitions, utilizing electronic medical record data. VE was evaluated with a test-negative case-control design using multivariate logistic regression with PCR-confirmed influenza as the outcome and vaccination status as the exposure, adjusting for age, enrolling site, illness onset date, race, days from onset to specimen collection, self-reported health, and self-reported hospitalizations.

**Results:** Of 3,524 adults hospitalized with ARI, 1,210 (34.3%) had an immunocompromising condition. IC adults were more likely to be vaccinated than non-IC (69.5% vs 65.2%), and less likely to have influenza (22% vs 27.8%). The mean age did not differ among IC and non-IC (61.4 vs 60.8 years old). The overall VE against influenza hospitalization, including immunocompetent adults, was 33% (95% CI, 21% to 44%). VE among IC vs non-IC adults was lower at 5% (−29% to 31%) vs. 41% (27% to 52%) (p<0.05 for interaction term).

**Conclusions:** VE in one influenza season was very low among IC individuals. Future efforts should include evaluation of VE among the different immunocompromising conditions and whether enhanced vaccines improve the suboptimal effectiveness among the immunocompromised.

## Introduction

The number of immunocompromised (IC) individuals has increased due to greater longevity of the population, increasing numbers of solid organ and stem cell transplants, advances in the treatment of hematologic and solid malignancies, increase in the number of individuals living with human immunodeficiency virus (HIV), and the use of steroids, immune-modulating agents, and other immunosuppressive drugs to treat autoimmune and inflammatory conditions [1, 2]. Immunosuppressive conditions are heterogeneous and the degree and type of immune deficiency caused by each one of these conditions vary, but a unifying consequence is an increased risk of many infectious diseases including influenza [3]. Influenza is a common cause of illness and death, with an estimated 140,000-810,000 influenza-associated hospitalizations and 12,000-61,000 influenza-associated deaths annually in the United States [4].

IC individuals are at higher risk for influenza-related complications, including increased frequency of hospitalization, ICU admission, longer duration of hospitalization, and death [5-10]. Influenza vaccination is the best available intervention for preventing these complications and annual influenza vaccination is recommended for IC individuals [11]. However, the data on protection afforded by influenza vaccines in IC adults are scarce. A recent study on cancer patients demonstrated a vaccine effectiveness (VE) of 20% against influenza hospitalization, as compared to 42% in the general population [12, 13]. Most studies of IC adults are small and evaluate immunogenicity as a surrogate of effectiveness [14]. These immunogenicity studies among various IC groups have demonstrated that antibody responses to inactivated influenza vaccines are suboptimal compared to those without immunosuppression [14, 15]. However, immune response to vaccine does not necessarily directly relate to vaccine effectiveness [16, 17]. Since the 2015-2016 influenza season, the Centers for Disease Control and Prevention (CDC)-funded U.S. Hospitalized Adult Influenza Vaccine Effectiveness Network (HAIVEN) has estimated influenza VE among adults hospitalized for acute respiratory infections.

Understanding influenza VE in IC individuals is crucial to the development of appropriate vaccination and public health policies. The purpose of this study was to evaluate influenza VE among hospitalized immunocompromised adults enrolled in the HAIVEN study during the 2017-2018 influenza season, when specific efforts were made to identify immunocompromised patients using case-definitions for immunocompromising conditions.

## Methods

### Study design and enrollment

The HAIVEN study is a multi-center, prospective, test-negative case-control study to determine an annual estimate of VE against influenza-associated hospitalizations among adults in the United States. Methods for the HAIVEN study have been described previously[18]. Briefly, adults ≥ 18 years of age with new or worsening cough or sputum production of ≤10 days’ duration and a respiratory specimen collected ≤ 10 days from illness onset and ≤ 72 hours after hospital admission at one of ten hospitals in Pennsylvania, Michigan, Tennessee, and Texas were eligible. Inclusion criteria included age ≥18 years, admission for an acute respiratory illness, or worsening of a chronic respiratory illness with a new or worsening cough. During the 2017-2018 influenza season, details on demographics, symptoms, influenza vaccination status, number of recent hospitalizations, and history of organ or stem cell transplant and, chemotherapy or radiation therapy in the preceding year were collected through the enrollment interview. Information about the clinical course and disease severity was obtained from electronic medical records (EMR). All international classification of diseases-10 diagnosis clinical modification (ICD-10-CM) codes and current procedural terminology (CPT) codes from all encounters in the 12 months before enrollment were obtained from the EMR and utilized to identify the high-risk conditions associated with an increased risk of serious influenza complications [11].

### Influenza case classification

Enrolled patients provided respiratory specimens for influenza testing by polymerase chain reaction (PCR). Specimens were either nasal and oropharyngeal swabs that were tested in research laboratories with CDC PCR protocols or clinical nasopharyngeal specimens tested by PCR in hospital laboratories provided they were collected within 10 days of illness onset and 72 hours of admission. Enrolled patients who tested positive for influenza were classified as cases and those who tested negative for all influenza types were controls.

### Influenza vaccination status

Self-reported current season influenza vaccination status was confirmed by medical record review, state immunization registry records, occupational health records, health insurance billing claims, and records from patients’ primary care providers. Information collected included date and route of administration and product name, manufacturer, and lot number. Self-reported vaccination was accepted if the patient provided a date and location for the vaccination. A participant was considered vaccinated if s/he received the 2017-18 influenza vaccine ≥ 14 days before illness onset. Because up to 14 days is required to mount an immune response to vaccination, those vaccinated 0 to 13 days before illness onset were excluded due to indeterminate vaccination status.

### Identification of immunocompromising conditions

All ICD-10-CM codes for all encounters and receipt of the biologic chemotherapeutic agents bortezomib, carfilzomib, daratumumab, dasatinib, gemtuzumab, and imatinib in the year before study enrollment were collected from EMR data. In the 2017-2018 influenza season the enrollment questionnaire asked if the participant received chemotherapy or radiation therapy for cancer in the 12 months before enrollment. Eight groups of immunocompromising conditions were defined: organ transplantation, stem cell transplantation, underlying immunodeficiency, connective tissue disorder, receipt of chemotherapy or radiation therapy, hematologic conditions, chronic steroid use, and HIV. The basis for the groups was a previously described algorithm for identifying patients with active immunosuppression utilizing ICD and CPT codes in a large database of patients with severe sepsis[19]. We slightly modified this algorithm in two aspects. For solid malignancies, we only included patients actively treated with chemotherapy or radiation to improve specificity of immunosuppression. We also included patients with chronic use of steroids. We considered the enrollment question on receipt of chemotherapy or radiation therapy as the gold standard and our data found that ICD-10-CM and CPT codes have low sensitivity to identify patients receiving chemotherapy or radiation therapy (Supplementary Table S1). Therefore, we identified patients with immunocompromising conditions based on ICD-10-CM codes listed (Table 1 and Supplementary Table S2), except for the receipt of chemotherapy or radiation therapy, which were determined from ICD-10-CM codes, or receipt of one of the biologic chemotherapeutic agents listed, or a positive answer to the enrollment question about the receipt of chemotherapy or radiation therapy.

**Table 1:** ICD-10 and CPT codes used to classify the immunocompromised group

| Immunocompromised Group | ICD-10 Codes |
| --- | --- |
| Organ Transplant | T86.1, T86.2, T86.3, T86.4, T86.81, T86.85, Z48.2, Z94.0, Z94.1, Z94.2, Z94.3, Z94.4, Z94.82, Z94.83 |
| Stem Cell Transplant | Z94.84, T86.0, T86.5 |
| Underlying Immunodeficiency | D80, D81.0, D81.1, D81.9, D82, D83, D84, D84.1, D84.8, D84.9, D89.8, D89.9 |
| Connective Tissue Disorder | M05, M06, M 08.0, M 08.2, M08.3, M08.4, L40.54, L40.59, M32, M30.0, M30.1, M30.2, M31.3, M33, M34, M34.0, M34.1, M34.9, M35.0, M35.9 |
| Chemotherapy/ Radiation Therapy | Z51.0, Z51.1 |
| Hematologic Conditions | C95.00, C95.10, D61.0, D61.2, D61.9, D70, D71, D72, D73.0, C81, C82, C83, C84, C85, C86, C88, C90, C91, C92, C93, C94, C96, D46 |
| Steroids | Z79.5, Z79.52 |
| HIV | B20, B97.35, O98.7, Z21 |
|  | CPT Codes |
| Chemotherapy | 96401-96417, 96420-96425, 96440-96450, G0498 |
| Radiation therapy | 77402-77412, G6003-G6014, 77385-77386, 77418, G6015-G6016, 77387, G6001-G6002, G6017, 77371-77372, 77373, 77778, 77770-77772, 77761-77763 |

CPT codes for chemotherapy and radiation therapy were collected at the Pennsylvania sites only.

The IC groups were mutually exclusive; therefore, if a participant had more than one IC condition, they were grouped hierarchically following the group order listed above. The hierarchical order of groups is shown in supplementary Figure S1 and was based on the authors’ expert opinion, to better identify active immunosuppressive conditions because we did not have data on the use of immunosuppressants other than steroids and biologicals in the dataset. For example, to identify patients with malignancies on active therapy, chemotherapy or radiation therapy preceded hematologic condition.

### Statistical analysis

Demographic and other characteristics of the IC and non-IC groups were compared using Pearson χ^2^ test or Fisher exact test for categorical variables and two-sample t-test for continuous variables.

VE was calculated by estimating the odds of influenza positivity among vaccinated patients compared to unvaccinated patients for the IC and non-IC groups using multivariate logistic regression using influenza positivity as the outcome and vaccination status as the exposure variable, with VE = (1 – adjusted odds ratio) × 100% [20].

In the primary analysis, we stratified the sample by immunocompromised status and estimated VE in each stratum:

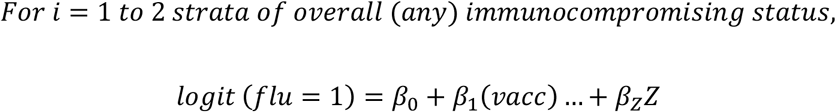

where

*flu* = 1 if PCR-confirmed flu case (of specific type/subtype); 0 otherwise

*vacc* = 1 if received vaccine ≥14 d prior to symptom onset; 0 otherwise

*Z =* vector of adjustment variables including age (continuous), enrollment site, race, days from illness onset to specimen collection, date of illness onset (categorized as pre-peak, peak, or post-peak influenza periods [18]), self-reported health status (poor/fair and good/very good/excellent) and self-reported number of hospitalizations

and with VE defined as

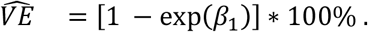

To test if VE differed by immunocompromised status, we regressed flu status on vaccination status, immunocompromised status, and the pairwise multiplicative interaction between vaccination status and immunocompromised status:

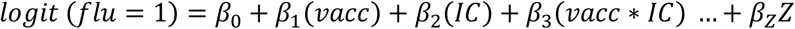

where variables are defined as above and

*IC* = 1 if immunocompromised (any immunocompromising condition); 0 otherwise

Effect modification of VE by immunocompromised status was assumed to be statistically significant if the test statistic for assessing if the coefficient for the interaction term, β_3_, differed from zero had a p value <0.05.

In secondary analyses, we stratified subjects by type-specific immunocompromised status and estimated VE within each stratum using a main effects model:

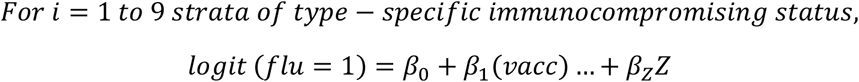

where variables are defined as above except for models for immunodeficiency and HIV subgroups in which

*Z =* vector of adjustment variables including age (continuous), enrollment site, race, days from illness onset to specimen collection, date of illness onset (categorized as pre-peak, peak, or post-peak influenza periods [18]), self-reported health status (poor/fair and good/very good/excellent) and self-reported number of hospitalizations

Because we did not specifically calculate sample sizes for this study, we did a post hoc power analysis based on the observed number of cases (n=900) and controls (2600), vaccination rate among controls (67%), power of 80%, and a significance level of 0.05. We determined a minimum detectable vaccine effectiveness of 20% in our overall study population during the 2017-2018 influenza season based on these assumptions.

Analyses were conducted with SAS version 9.4 software. Statistical significance was defined as a p-value < 0.05 or a 95% confidence interval (CI) excluding the null value. We interpreted differences in VE estimates by IC vs non-IC subgroups, considering p-value <0.15 as statistically significant which is in line with guidance for interpreting interaction between two dichotomous variables when effect size is expected to be moderate to high[21, 22]. The study protocol was approved by the research ethics boards at the participating institutions.

## Results

A total of 4,108 hospitalized adults were enrolled in HAIVEN in the 2017-2018 influenza season. Of these, 584 were excluded because of enrollment earlier or later than the period of influenza circulation in the community (n=259), missing vaccination status (n=201), missing number of self-reported past year hospitalizations (n=59), and other reasons (n=65) (Figure 1). In the resulting dataset (n=3,524), 1,210 (34.3%) adults were identified as having an immunocompromising condition: organ transplant (n=144, 11.9%); stem cell transplant (n=28, 2.3%); underlying immunodeficiency (n=49, 4.0%); connective tissue and rheumatologic disease (n=130, 10.7%); chemotherapy and radiation therapy (n=242, 20%); hematologic condition (n=175, 14.5%); chronic steroid use (n=397, 32.8%) and HIV (n=45, 3.7%).

**Figure 1:**
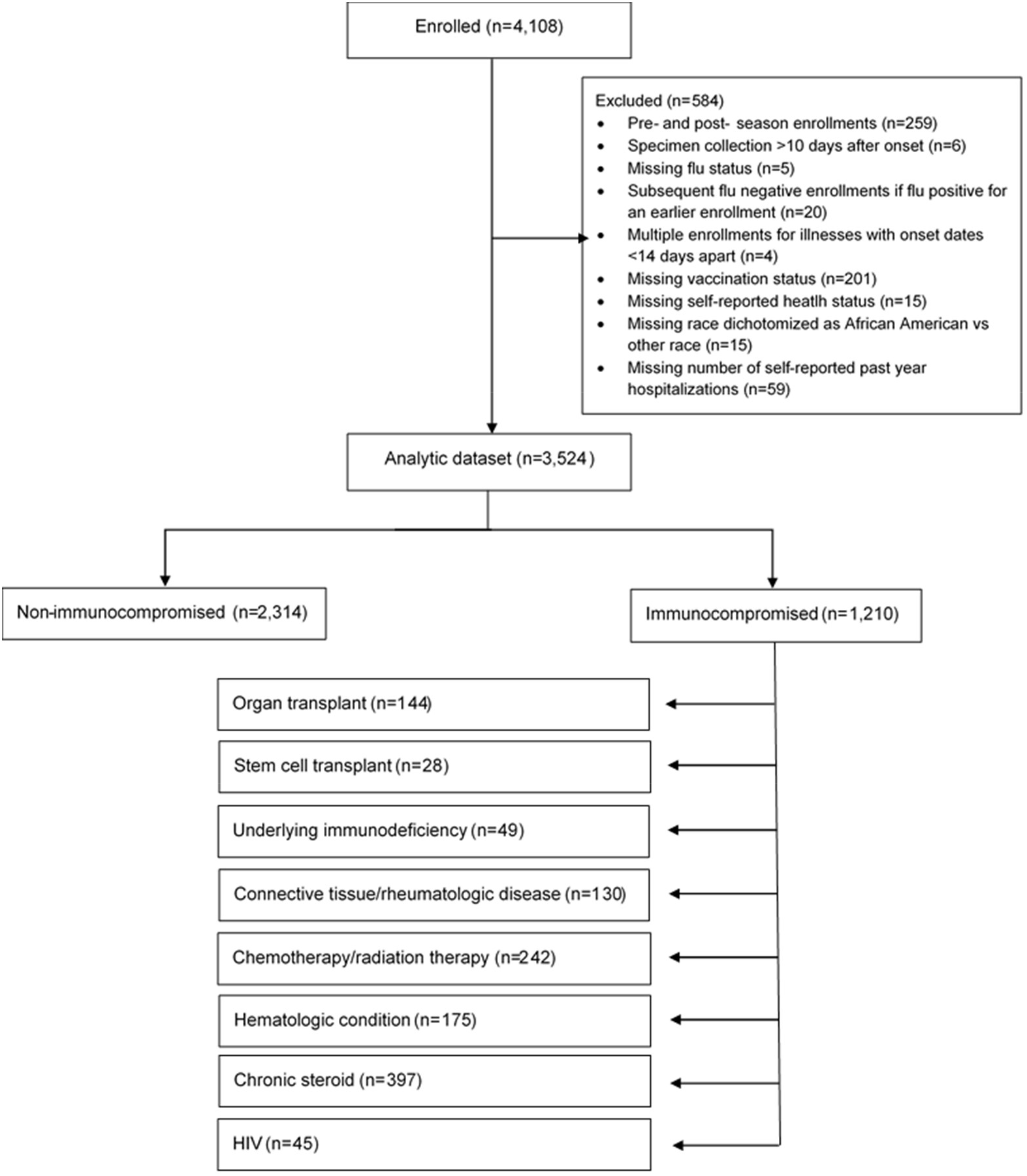
US Hospitalized Adult Influenza Vaccine Effectiveness Network (HAIVEN) study population, 2017–2018. Immunocompromised groups were mutually exclusive and followed the order listed here. As an example, if an individual had an ICD-10 code that classified him/her as an organ transplant, this individual was grouped in this category, even if he/she also had an ICD-10 for chemotherapy.

Overall, participants were more likely to be female (56.9%) and white (62.2%). Mean age was 61 (SD 17.1) years, 66.7% were vaccinated, 25.8% had influenza, and 84.2% had ≥ 3 high-risk conditions (Table 2). The IC and non-IC groups differed for several characteristics. IC participants were significantly more likely than non-IC to be of white race (67.9% vs 59.3%, p<0.001), have a lower BMI (30.1 vs 31.2, p=0.003), be vaccinated (60.2% vs 54.6%, p=0.002), have a longer length of stay (4 vs 3 days, p<0.001), have ≥ 3 high-risk conditions (94.2% vs 79%, p<0.001), have had ≥ 4 hospitalizations in the previous year (25.5% vs 19.1%, p<0.001), and present earlier in the pre-peak period (42.1% vs 37.4%, p=0.02). IC participants were significantly less likely than non-IC participants to test positive for influenza (22% vs 27.8%, p<0.001) and to self-report their health as fair or poor (45% vs 53.6%, p<0.001) (Table 2).

**Table 2:** Patient characteristics overall by immunocompromising condition, US Hospitalized Adult Influenza Vaccine Effectiveness (HAIVEN) study, 2017-2018 (n=3,524)

|  | Total<br>(n= 3,524) | Non-Immunocompromised<br>(n=2,314) | Immunocompromised<br>(n=1,210) | p-value |
| --- | --- | --- | --- | --- |
| Enrollment site, n (%) |  |  |  |  |
| Michigan | 943 (26.8) | 714 (30.9) | 229 (18.9) | **<0.001** |
| Pennsylvania | 834 (23.7) | 571 (24.7) | 263 (21.7) |  |
| Tennessee | 589 (16.7) | 369 (16.0) | 220 (18.2) |  |
| Texas | 1158 (32.9) | 660 (28.5) | 498 (41.2) |  |
| Female, n (%) | 2004 (56.9) | 1317 (56.9) | 687 (56.8) | 0.94 |
| Age group, n (%) |  |  |  |  |
| 18-49 | 790 (22.4) | 534 (23.1) | 256 (21.2) | 0.37 |
| 50-64 | 1173 (33.3) | 753 (32.5) | 420 (34.7) |  |
| 65-74 | 798 (22.6) | 517 (22.3) | 281 (23.2) |  |
| 75+ | 763 (21.7) | 510 (22.0) | 253 (20.9) |  |
| Age, mean ± SD | 61.0 ± 17.1 | 60.8 ± 17.7 | 61.4 ± 15.9 | 0.29 |
| Race, n (%) |  |  |  |  |
| White, non-Hispanic | 2193 (62.2) | 1371 (59.3) | 822 (67.9) | **<0.001** |
| Non-white | 1331 (37.8) | 943 (40.8) | 388 (32.1) |  |
| BMI, mean ± SD | 30.8 ± 9.5 | 31.2 ± 9.8 | 30.1 ± 8.9 | **0.001** |
| Any flu, n (%) |  |  |  |  |
| Negative | 2614 (74.2) | 1670 (72.2) | 944 (78.0) | **<0.001** |
| Positive | 910 (25.8) | 644 (27.8) | 266 (22.0) |  |
| Documented vaccination, n (%) |  |  |  |  |
| No | 1174 (33.3) | 805 (34.8) | 369 (30.5) | **0.01** |
| Yes | 2350 (66.7) | 1509 (65.2) | 841 (69.5) |  |
| Length of stay, median (IQR) | 3.0 (4.0) | 3.0 (3.0) | 4.0 (4.0) | **<0.001** |
| Influenza-like illness (ILI) |  |  |  |  |
| symptoms, n (%) |  |  |  |  |
| No | 1208 (34.3) | 797 (34.4) | 411 (34.0) | 0.78 |
| Yes | 2316 (65.7) | 1517 (65.6) | 799 (66.0) |  |
| Number of high-risk conditions, |  |  |  |  |
| n (%) |  |  |  |  |
| No high-risk conditions | 162 (4.6) | 141 (6.1) | 21 (1.7) | **<0.001** |
| 1-2 high-risk conditions | 394 (11.2) | 345 (14.9) | 49 (4.1) |  |
| ≥3 high-risk conditions | 2968 (84.2) | 1828 (79.0) | 1140 (94.2) |  |
| Self-reported hospitalizations in |  |  |  |  |
| the prior year, n (%) |  |  |  |  |
| 0-3 hospitalizations | 2773 (78.7) | 1871 (80.9) | 902 (74.6) | **<0.001** |
| ≥4 hospitalizations | 751 (21.3) | 443 (19.1) | 308 (25.5) |  |
| Interval from illness onset and |  |  |  |  |
| specimen collection, n (%) |  |  |  |  |
| 0-1 days | 693 (19.7) | 435 (18.8) | 258 (21.3) | 0.14 |
| 2-4 days | 1621 (46.0) | 1065 (46.0) | 556 (46.0) |  |
| 5-10 days | 1210 (34.3) | 814 (35.2) | 396 (32.7) |  |
| Onset date |  |  |  |  |
| Pre-peak | 1374 (39.0) | 865 (37.4) | 509 (42.1) | **0.02** |
| Peak | 844 (24.0) | 559 (24.2) | 285 (23.6) |  |
| Post-peak | 1306 (37.1) | 890 (38.5) | 416 (34.4) |  |
| Self-reported health status, n |  |  |  |  |
| (%) |  |  |  |  |
| Excellent/ very good/ good | 1739 (49.4) | 1074 (46.4) | 665 (55.0) | **<0.001** |
| Fair/ poor | 1785 (50.7) | 1240 (53.6) | 545 (45.0) |  |

There were 266 influenza cases in the IC adults and 644 influenza cases in non-IC adults. Most influenza infections were caused by influenza A, and 530 (78.8%) were A(H3N2) viruses. Of 238 influenza B infections, 200 (84%) were due to B Yamagata lineage viruses (Figure 2).

**Figure 2:**
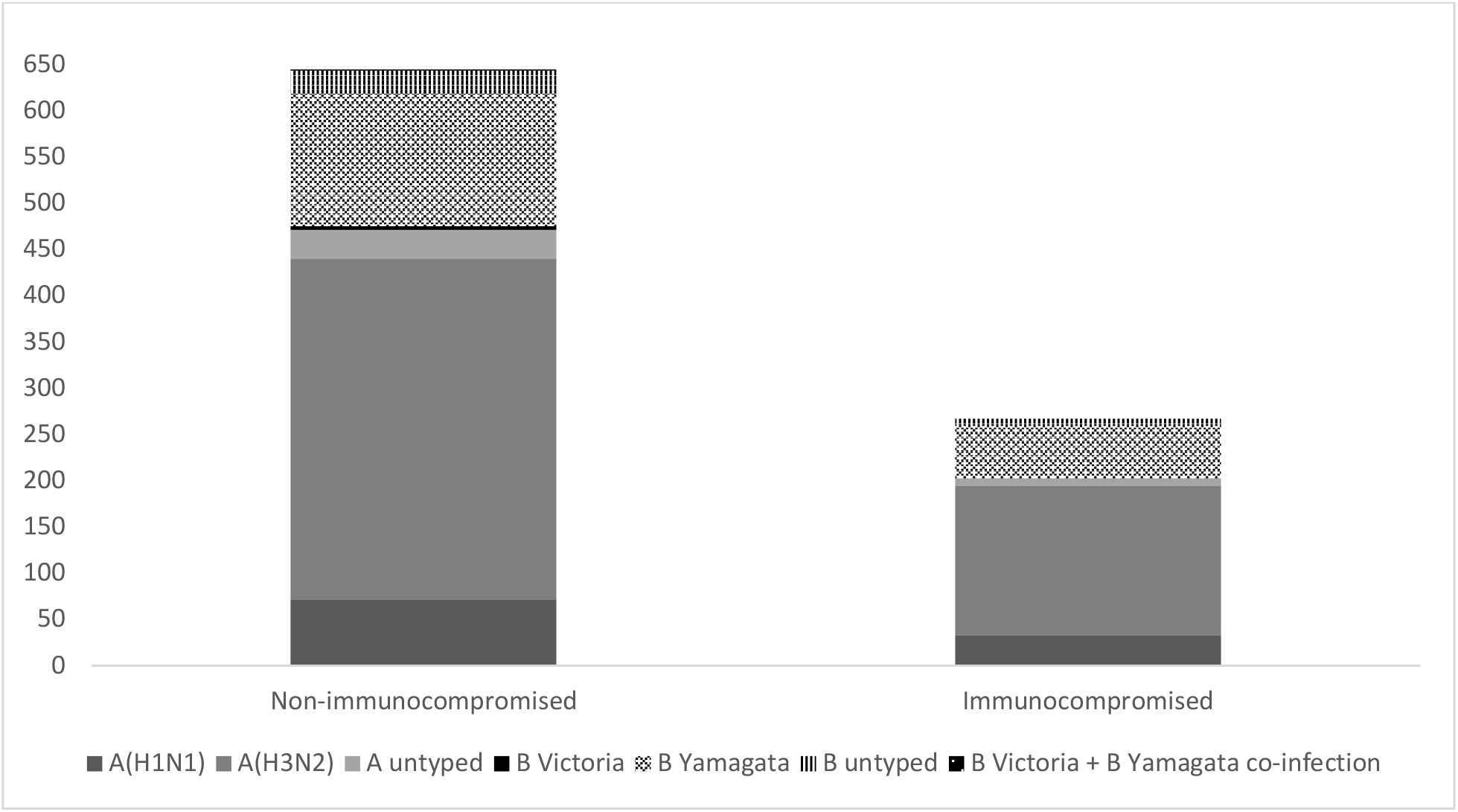
Influenza virus type/subtype and lineage in the non-immunocompromised and immunocompromised groups.

The patients in the 8 immunocompromised groups differed in sex, enrollment site, age/age group, race, BMI, influenza status, documented influenza vaccination, number of high-risk conditions, and self-reported health status, but not in the number of hospitalizations in the previous year, interval from illness onset to specimen collection, and date of illness onset (Supplementary Table S3).

Overall, vaccination was 33% (95% CI, 21% to 44%) effective in preventing hospitalization. Among IC adults, VE was 5% and not significant (95% CI, −29% to 31%). VE in non-IC adults was 41% (95% CI, 27% to 52%) (p<0.05 for interaction term) (Table 3). VE for the different immunocompromised conditions varied widely, from −73% for individuals with underlying immunodeficiency to 84% for stem cell transplant; however, this study was not powered to look at these subgroups and the confidence intervals varied widely (Supplementary figure S2).

**Table 3:** Influenza vaccine effectiveness for prevention of influenza A or B-associated hospitalizations in immunocompromised and non-immunocompromised adults, US Hospitalized Adult Influenza Vaccine Effectiveness (HAIVEN) study, 2017-2018

|  | N Influenza Cases (% vaccinated) | Unadjusted VE, % (95% CI) | Adjusted VE, % (95% CI)* |
| --- | --- | --- | --- |
| All, n=3524 | 910 (67) | 28 (16, 38) | 33 (21, 44) |
| Non-immunocompromised, n=2314 | 644 (65) | 36 (23, 47) | 41 (27, 52) |
| Immunocompromised, n=1210 | 266 (70) | -0.3 (-35, 25) | 5 (-29, 31) |
| Non-immunocompromised, influenza A | 471 (65) | 31 (15, 44) | 31 (14, 46) |
| Immunocompromised, influenza A | 202 (70) | 1 (-37, 29) | 4 (-36, 32) |
| Non-immunocompromised, H1N1 | 71 (65) | 57 (32, 74) | 52 (19, 71) |
| Immunocompromised, H1N1 | 33 (70) | 60 (17, 80) | 51 (-2, 77) |
| Non-immunocompromised, H3N2 | 369 (65) | 21 (0, 37) | 23 (0, 40) |
| Immunocompromised, H3N2 | 161 (70) | -28 (-87, 12) | -18 (-75, 20) |
| Non-immunocompromised, influenza B | 173 (65) | 34 (10, 52) | 45 (22, 61) |
| Immunocompromised, influenza B | 65 (70) | 1 (-70, 43) | 13 (-52, 51) |

\* Adjusted for enrolling site, onset date (pre-peak, peak, post-peak), age, race, days from illness onset to specimen collection (0-1, 2-4, 5-10 days), self-reported health (poor/fair, good/very good/excellent), and self-reported hospitalizations.

## Discussion

During the high-severity 2017-2018 US influenza season, we found that influenza vaccination reduced the risk of influenza-associated hospitalization among adults by 33%. Overall, VE during the 2017-2018 season was lower than that estimated in previous seasons in this network (42-54%) [18, 23]. The fact that influenza A(H3N2) viruses circulating in 2017-2018 were antigenically different from the vaccine H3N2 strain because of suspected egg-adapted glycosylation in the antigenic epitopes of the vaccines may be responsible for the lower VE [24]. As compared with VE in non-IC adults (41%), VE in IC adults was significantly lower (5%) during this season. This lower VE among IC adults is unlikely to be an artifact, because the findings are consistent with the immunogenicity studies of inactivated influenza vaccines (IIVs) that have demonstrated significantly reduced humoral immune responses to standard IIVs in immunosuppressed patients with HIV, organ transplants, cancer, and those receiving immunosuppressants [14]. In this network, influenza vaccination rate among controls was greater in the IC (60%) than in the non-IC group (54%), which is consistent with national US data in the insured population [25]. The higher vaccination rate among IC may be due to more frequent healthcare encounters and closer monitoring among IC patients offering more opportunities to vaccinate, or a heightened perception of risk for influenza complications by providers, leading to increased willingness to recommend influenza vaccine, and by patients, leading to greater willingness to receive vaccination.

Limited data exist on the prevention of influenza infection on immunocompromised adults by vaccination. Most studies have focused on the measurement of humoral antibody response among patients with particular immunocompromising conditions and have reported significantly reduced humoral immune responses [15, 26, 27]. However, this approach disregards the relationship between clinical outcomes and immune response, the levels of antibody titers from previous immunizations that may cause overestimations of response, nor does it consider the role of cell-mediated immune response to vaccination in the prevention of influenza infection. While studies of high dose influenza vaccine have demonstrated improved antibody responses in adult organ transplant recipients and improved antibody responses and outcomes in adults older than 65 years of age as compared to standard dose vaccine, it is unknown if enhanced vaccine options, such as high dose and adjuvanted vaccines, could improve VE in immunocompromised groups [27-30]. Increasing the evidence base for informing the use of enhanced influenza vaccines in immunosuppressed populations is necessary for determining if these interventions might offer added value to standard influenza vaccines and potentially contribute to improving efficacy of these vaccines.

A primary challenge in the study of influenza VE in IC individuals is the definition of immunocompromise. Immunocompromising conditions are heterogenous and the degree of immunosuppression among groups is challenging to quantify. Additionally, within a defined IC group, differences in the degree of immunosuppression are difficult to assess, based on clinical records. We considered ∼34% of the adults hospitalized with an acute respiratory illness during the influenza season as being immunocompromised by pre-defining groups of immunocompromising conditions that were identified by ICD-10-CM codes for all medical encounters in the preceding year. To complement our case definition, we also analyzed the addition of CPT codes for chemotherapy administration, chemotherapeutic drugs recorded in the EMR, and a question at the time of enrollment about the receipt of chemotherapy or radiation therapy in the preceding 12 months. Although we did not collect other immunosuppressant and biological data, we identified a similar proportion of IC adults among those hospitalized with acute respiratory illness as identified in the study by Patel et al. that utilized MarketScan data to estimate the prevalence of immunosuppressive conditions and risk for acute respiratory illnesses [25].

Findings of this study should be interpreted in the context of several limitations. Although we used an objective and systematic mechanism to identify the different IC groups, our identification of the immunocompromising groups accounts for only a rough measure of immunosuppression. We did not consider the presence of more than one immunocompromising condition, and we were unable to evaluate the effect of timing of vaccination in relation to the immunosuppression. We were unable to evaluate VE among different IC because of inadequate sample sizes. A study with a greater number of IC adults that allows for analyses of subgroups, virus subtypes, and different vaccine formulations is needed for definitive conclusions. Our study is also limited to a single season when vaccine was mismatched to the circulating A/H3N2 viruses and thus may not be applicable to other influenza viruses. Data are also from 4 U.S. sites and may not be generalizable.

Our study’s strengths include the use of a standardized protocol with symptom-based eligibility and comprehensive PCR testing to identify influenza cases and controls, a test-negative case control design and, recruitment in geographically diverse areas. Furthermore, our study shows that immunocompromising conditions can be identified based on EMR data, without the need for cumbersome medication reviews.

Proper identification of IC groups in future VE studies will have implications for public policy development, such as a recommendation for a different vaccine formulation for IC groups, or a consideration for chemoprophylaxis for those with immunocompromise.

Vaccine effectiveness against influenza was not significant among hospitalized immunocompromised patients. In light of our findings, decreasing the burden of influenza in IC individuals may be less dependent on improving their vaccine coverage than on improving vaccination rates of close contacts of the immunocompromised individual, thereby creating a circle of protection around an IC individual. Mathematical modeling has shown that even small improvements in VE and vaccine coverage are associated with substantial reductions in influenza burden [31].

## Data Availability

Data may be made available after completion of the study.

## Notes

### HAIVEN Study Investigators

University of Pittsburgh Medical Center, Pennsylvania: Richard Zimmerman, Donald Middleton, Fernanda Silveira, Kailey Hughes, Heather Eng, Theresa Sax, Sean Saul, Charles Rinaldo, Balasubramani Goundappa, Mary Patricia Nowalk, Lori Steiffel, John Williams, Monika Johnson. Baylor Scott and White Health, Texas: Manjusha Gaglani, Kempapura Murthy, Tresa McNeal, Shekar Ghamande, Victor Escobedo, Anne Robertson, Lydia Clipper, Arundhati Rao, Kevin Chang, Marcus Volz, Kimberly Walker, Alejandro Arroliga. University of Michigan, Michigan: Arnold Monto, Emily Martin, Ryan Malosh, Joshua Petrie, Adam Lauring, Caroline Cheng, Hannah Segaloff, E. J. McSpadden, Emileigh Johnson, Rachel Truscon. Henry Ford Health System, Michigan: Lois Lamerato, Susan Davis, Marcus Zervos. Vanderbilt University Medical Center, Tennessee: H. Keipp Talbot, Dayna Wyatt, Yuwei Zhu, Zhouwen Liu, Rendie McHenry, Natasha Halasa, Sandra Alvarez Calvillo, Stephanie Longmire, Laura Stewart. CDC: Jill Ferdinands, Alicia Fry, Elif Alyanak, Emily Smith, Courtney Strickland, Sarah Spencer, Brendan Flannery, Jessie Chung, Xiyan Xu, Stephen Lindstrom, LaShondra Berman, Wendy Sessions, Rebecca Kondor, Manish Patel.

### Disclaimer

The findings and conclusions in this report are those of the authors and do not necessarily represent the official position of CDC.

