## Supplementary material for "Effectiveness of Influenza Vaccine for Preventing Laboratory-Confirmed Influenza Hospitalizations in Immunocompromised Adults"

**Table S1:** Sensitivity, specificity, positive predictive value, and negative predictive value for CPT codes and ICD-10 codes for chemotherapy or radiation therapy with the enrollment question on receipt of chemotherapy or radiation therapy for cancer in the 12 months before enrollment as the gold standard

| | Sensitivity | Specificity | Positive Predictive Value | Negative Predictive Value |
|---|---|---|---|---|
| CPT code for chemotherapy or radiation therapy[1] | 3 | 99.5 | 43 | 89.6 |
| ICD-10 code for chemotherapy or radiation therapy | 2.4 | 99.9 | 87.5 | 91.7 |

Data presented as %
1 Data from Pennsylvania site hospitals only

**Table S2:** ICD-10 and CPT codes used to classify the immunocompromised groups

| Immunocompromising Condition | ICD-10 code |
|---|---|
| ***Organ transplant*** | |
| Complications of kidney transplant | T86.1 |
| Complications of heart transplant | T86.2 |
| Complications of heart-lung transplant | T86.3 |
| Complications of liver transplant | T86.4 |
| Complications of lung transplant | T86.81 |
| Complications of intestine transplant | T86.85 |
| Encounter for aftercare following organ transplant | Z48.2 |
| Kidney transplant status | Z94.0 |
| Heart transplant status | Z94.1 |
| Lung transplant status | Z94.2 |
| Heart and lung transplant status | Z94.3 |
| Liver transplant status | Z94.4 |
| Intestine transplant status | Z94.82 |
| Pancreas transplant status | Z94.83 |
| ***Stem cell transplant*** | |
| Stem cells transplant status | Z94.84 |
| Complications of bone marrow transplant | T86.0 |
| Complications of stem cell transplant | T86.5 |
| ***Underlying immunodeficiency*** | |
| Immunodeficiency with predominantly antibody defects | D80 |
| Severe combined immunodeficiency [SCID] with reticular dysgenesis | D81.0 |
| Severe combined immunodeficiency [SCID] with low T- and B-cell numbers | D81.1 |
| Severe combined immunodeficiency [SCID] with low or normal B-cell numbers | D81.2 |
| Combined immunodeficiency, unspecified | D81.9 |
| Immunodeficiency associated with other major defects | D82 |
| Common variable immunodeficiency | D83 |
| Other immunodeficiencies | D84 |
| Defects in the complement system | D84.1 |

| | |
|---|---|
| Other specified immunodeficiencies | D84.8 |
| Immunodeficiency, unspecified | D84.9 |
| Other specified disorders involving the immune mechanism, not elsewhere classified | D89.8 |
| Disorder involving the immune mechanism, unspecified | D89.9 |
| ***Connective tissue disorder*** | |
| Rheumatoid arthritis with rheumatoid factor | M05 |
| Other rheumatoid arthritis | M06 |
| Unspecified juvenile rheumatoid arthritis | M08.0 |
| Juvenile rheumatoid arthritis with systemic onset | M08.2 |
| Juvenile rheumatoid polyarthritis (seronegative) | M08.3 |
| Pauciarticular juvenile rheumatoid arthritis | M08.4 |
| Psoriatic juvenile arthropathy | L40.54 |
| Other psoriatic arthropathy | L40.59 |
| Systemic lupus erythematosus | M32 |
| Polyarteritis nodosa | M30.0 |
| Polyarteritis with lung involvement [Churg-Strauss] | M30.1 |
| Juvenile polyarteritis | M30.2 |
| Wegener's granulomatosis | M31.3 |
| Dermatopolymyositis | M33 |
| Systemic sclerosis [scleroderma] | M34 |
| Progressive systemic sclerosis | M34.0 |
| CR(E)ST syndrome | M34.1 |
| Systemic sclerosis, unspecified | M34.9 |
| Sicca syndrome [Sjogren] | M35.0 |
| Systemic involvement of connective tissue, unspecified | M35.9 |
| ***Chemotherapy or radiation therapy*** | |
| Encounter for antineoplastic radiation therapy | Z51.0 |
| Encounter for antineoplastic chemotherapy and immunotherapy | Z51.1 |
| ***Hematologic conditions*** | |
| Acute leukemia of unspecified cell type not having achieved remission | C95.00 |
| Chronic leukemia of unspecified cell type not having achieved remission | C95.10 |

| | |
|---|---|
| Constitutional aplastic anemia | D61.0 |
| Idiopathic aplastic anemia | D61.2 |
| Aplastic anemia, unspecified | D61.9 |
| Neutropenia | D70 |
| Functional disorders of polymorphonuclear neutrophils | D71 |
| Other disorders of white blood cells | D72 |
| Hyposplenism | D73.0 |
| Hodgkin lymphoma | C81 |
| Follicular lymphoma | C82 |
| Non-follicular lymphoma | C83 |
| Mature T/NK-cell lymphoma | C84 |
| Other specified and unspecified types of non-Hodgkin lymphoma | C85 |
| Other specified types of T/NK-cell lymphoma | C86 |
| Malignant immunoproliferative diseases and certain other B-cell lymphomas | C88 |
| Multiple myeloma and malignant plasma cell neoplasms | C90 |
| Lymphoid leukemia | C91 |
| Myeloid leukemia | C92 |
| Monocytic leukemia | C93 |
| Other leukemias of specified cell type | C94 |
| Other and unspecified malignant neoplasms of lymphoid, hematopoietic and related tissue | C96 |
| Myelodysplastic syndromes | D46 |
| ***Chronic use of steroids*** | |
| Long term (current) use of steroids | Z79.5 |
| Long term (current) use of systemic steroids | Z79.52 |
| ***HIV*** | |
| Human immunodeficiency virus [HIV] disease | B20 |
| Human immunodeficiency virus, type 2 [HIV 2] as the cause of diseases classified elsewhere | B97.35 |
| Human immunodeficiency virus [HIV] disease complicating pregnancy, childbirth and the puerperium | O98.7 |
| Asymptomatic human immunodeficiency virus [HIV] infection status | Z21 |
| | **CPT code** |

| | |
|---|---|
| ***Chemotherapy*** | 96401-96417 |
| | 96420-96425 |
| | 96440-96450 |
| | G0498 |
| ***Radiation therapy*** | |
| External Beam Radiation Therapy | 77402-77412<br>G6003-G6014 |
| Intensity Modulated Radiation Therapy | 77385-77386<br>77418<br>G6015-G6016 |
| Image-guided Radiation Therapy | 77387<br>G6001-G6002<br>G6017 |
| Stereotactic Radiosurgery | 77371-77372 |
| Stereotactic Body Radiation Therapy | 77373 |
| Brachytherapy | 77778<br>77770-77772 |
| Intracavitary Radiation Therapy | 77761-77763 |

**Table S3:** Patient characteristics overall by type of immunocompromising conditions, US Hospitalized Adult Influenza Vaccine Effectiveness (HAIVEN) study, 2017-2018 (n=1,210)

| | Organ Transplant (n=144) | Stem Cell Transplant (n=28) | Underlying Immunodeficiency (n=49) | Connective Tissue/ Rheumatologic Disease (n=130) | Chemo/ Radiation Therapy (n=242) | Hematologic Condition (n=175) | Chronic Steroid Use (n=397) | HIV (n=45) | p-value |
|---|---|---|---|---|---|---|---|---|---|
| Female, n (%) | 55 (38.2) | 11 (39.3) | 22 (44.9) | 104 (80.0) | 117 (48.4) | 105 (60.0) | 259 (65.2) | 14 (31.1) | **<0.001** |
| Enrollment site, n (%) | | | | | | | | | |
| Michigan | 43 (29.9) | 12 (42.9) | 14 (28.6) | 29 (22.3) | 55 (22.7) | 31 (17.7) | 31 (7.8) | 14 (31.1) | **<0.001** |
| Pennsylvania | 45 (31.3) | 6 (21.4) | 8 (16.3) | 31 (23.9) | 68 (28.1) | 35 (20.0) | 66 (16.6) | 4 (8.9) | |
| Tennessee | 33 (22.9) | 8 (28.6) | 11 (22.5) | 29 (22.3) | 40 (16.5) | 59 (33.7) | 31 (7.8) | 9 (20.0) | |
| Texas | 23 (16.0) | 2 (7.1) | 16 (32.7) | 41 (31.5) | 79 (32.6) | 50 (28.6) | 269 (67.8) | 18 (40.0) | |
| Age Group, n (%) | | | | | | | | | |
| 18-49 | 37 (25.7) | 5 (17.9) | 13 (26.5) | 31 (23.9) | 42 (17.4) | 43 (24.6) | 65 (16.4) | 20 (44.4) | **<0.001** |
| 50-64 | 58 (40.3) | 12 (42.9) | 13 (26.5) | 46 (35.4) | 75 (31.0) | 69 (39.4) | 126 (31.7) | 21 (46.7) | |
| 64-74 | 40 (27.8) | 9 (32.1) | 14 (28.6) | 29 (22.3) | 71 (29.3) | 25 (14.3) | 89 (22.4) | 4 (8.9) | |
| 75+ | 9 (6.3) | 2 (7.1) | 9 (18.4) | 24 (18.5) | 54 (22.3) | 38 (21.7) | 117 (29.5) | 0 (0.0) | |
| Age, mean ± SD | 57.5 ± 13.5 | 60.1 ± 13.3 | 60.8 ± 17.9 | 60.4 ± 15.8 | 63.2 ± 14.7 | 59.5 ± 18.1 | 64.4 ± 15.7 | 49.8 ± 10.8 | **<0.001** |
| Race, n (%) | | | | | | | | | |
| White, non-Hispanic | 98 (68.1) | 24 (85.7) | 41 (83.7) | 78 (60.0) | 179 (74.0) | 113 (64.6) | 268 (67.5) | 21 (46.7) | **<0.001** |
| Non-White | 46 (31.9) | 4 (14.3) | 8 (16.3) | 52 (40.0) | 63 (26.0) | 62 (35.4) | 129 (32.5) | 24 (53.3) | |
| BMI, mean ± SD | 28.2 ± 6.3 | 27.1 ± 6.2 | 29.4 ± 8.3 | 29.9 ± 8.3 | 28.4 ± 7.3 | 30.8 ± 8.9 | 31.9 ± 10.3 | 29.7 ± 12.2 | **<0.001** |
| Any Flu, n (%) | 37 (25.7) | 13 (46.4) | 12 (24.5) | 31 (23.9) | 50 (20.7) | 35 (20.0) | 75 (18.9) | 13 (28.9) | **0.03** |
| Documented vaccination, n (%) | 109 (75.7) | 20 (71.4) | 33 (67.4) | 87 (66.9) | 145 (59.9) | 107 (61.1) | 306 (77.1) | 34 (75.6) | **<0.001** |
| Length of Stay, median (IQR) | 4.0 (3.5) | 4.0 (4.0) | 4.0 (4.0) | 4.0 (5.0) | 3.0 (4.0) | 4.0 (5.0) | 4.0 (3.0) | 3.0 (3.0) | 0.08 |
| Number of high-risk conditions, n (%) | | | | | | | | | |
| None | 2 (1.4) | 0 (0.0) | 0 (0.0) | 3 (2.3) | 6 (2.5) | 4 (2.3) | 5 (1.3) | 1 (2.2) | **0.03** |
| 1-2 | 1 (0.7) | 0 (0.0) | 1 (2.0) | 5 (3.9) | 10 (4.1) | 17 (9.7) | 12 (3.0) | 3 (6.7) | |

| | | | | | | | | | |
|---|---|---|---|---|---|---|---|---|---|
| ≥3 | 141 (97.9) | 28 (100.0) | 48 (98.0) | 122 (93.9) | 226 (93.4) | 154 (88.0) | 380 (95.7) | 41 (91.1) | |
| Self-reported hospitalizations in the prior year, n (%) | | | | | | | | | |
| 0-3 hospitalizations | 105 (72.9) | 17 (60.7) | 38 (77.6) | 101 (77.7) | 171 (70.7) | 137 (78.3) | 297 (74.8) | 36 (80.0) | 0.34 |
| ≥4 hospitalizations | 39 (27.1) | 11 (39.3) | 11 (22.5) | 29 (22.3) | 71 (29.3) | 38 (21.7) | 100 (25.2) | 9 (20.0) | |
| Interval from illness onset and specimen collection, n (%) | | | | | | | | | |
| 0-1 days | 29 (20.1) | 2 (7.1) | 10 (20.4) | 25 (19.2) | 57 (23.6) | 32 (18.3) | 95 (23.9) | 8 (17.8) | 0.29 |
| 2-4 days | 57 (39.6) | 19 (67.9) | 23 (46.9) | 64 (49.2) | 106 (43.8) | 79 (45.1) | 184 (46.4) | 24 (53.3) | |
| 5-10 days | 58 (40.3) | 7 (25.0) | 16 (32.7) | 41 (31.5) | 79 (32.6) | 64 (36.6) | 118 (29.7) | 13 (28.9) | |
| Onset date | | | | | | | | | |
| Pre-peak | 53 (36.8) | 12 (42.9) | 18 (36.7) | 48 (36.9) | 100 (41.3) | 80 (45.7) | 180 (45.3) | 18 (40.0) | 0.13 |
| Peak | 46 (31.9) | 6 (21.4) | 9 (18.4) | 27 (20.8) | 58 (24.0) | 30 (17.1) | 99 (24.9) | 10 (22.2) | |
| Post-peak | 45 (31.3) | 10 (35.7) | 22 (44.9) | 55 (42.3) | 84 (34.7) | 65 (37.1) | 118 (29.7) | 17 (37.8) | |
| Self-reported health status, n (%) | | | | | | | | | |
| Excellent/ very good/ good | 63 (43.8) | 13 (46.4) | 29 (59.2) | 83 (63.9) | 121 (50.0) | 89 (50.9) | 239 (60.2) | 28 (62.2) | **0.003** |
| Fair/ poor | 81 (56.3) | 15 (53.6) | 20 (40.8) | 47 (36.2) | 121 (50.0) | 86 (49.1) | 158 (39.8) | 17 (37.8) | |

**Figure S1: Algorithm for identification of immunocompromised groups**

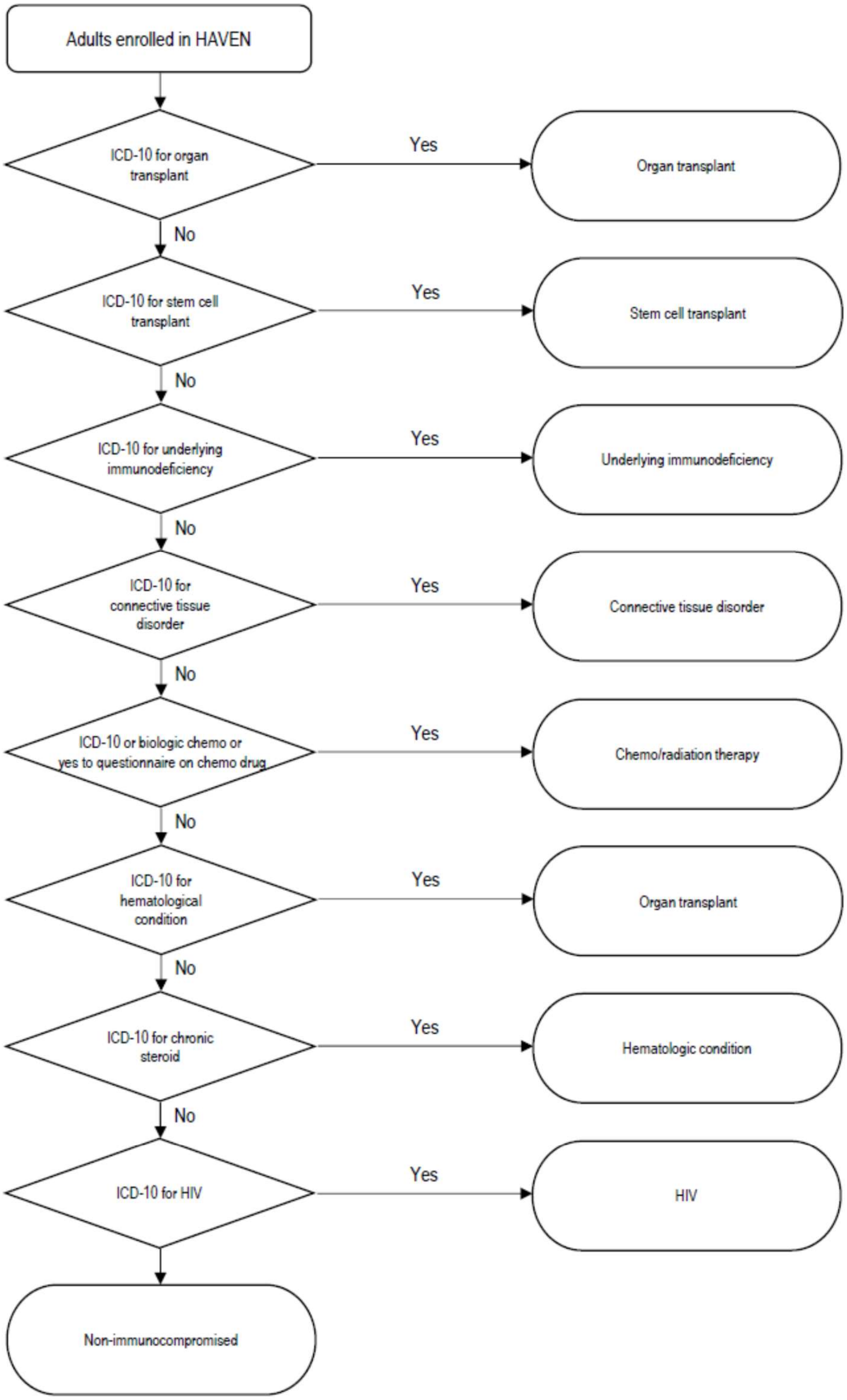

**Figure S2:** Adjusted vaccine effectiveness against influenza A and B among patients with specific immunocompromising conditions, HAIVEN study, 2017-2018

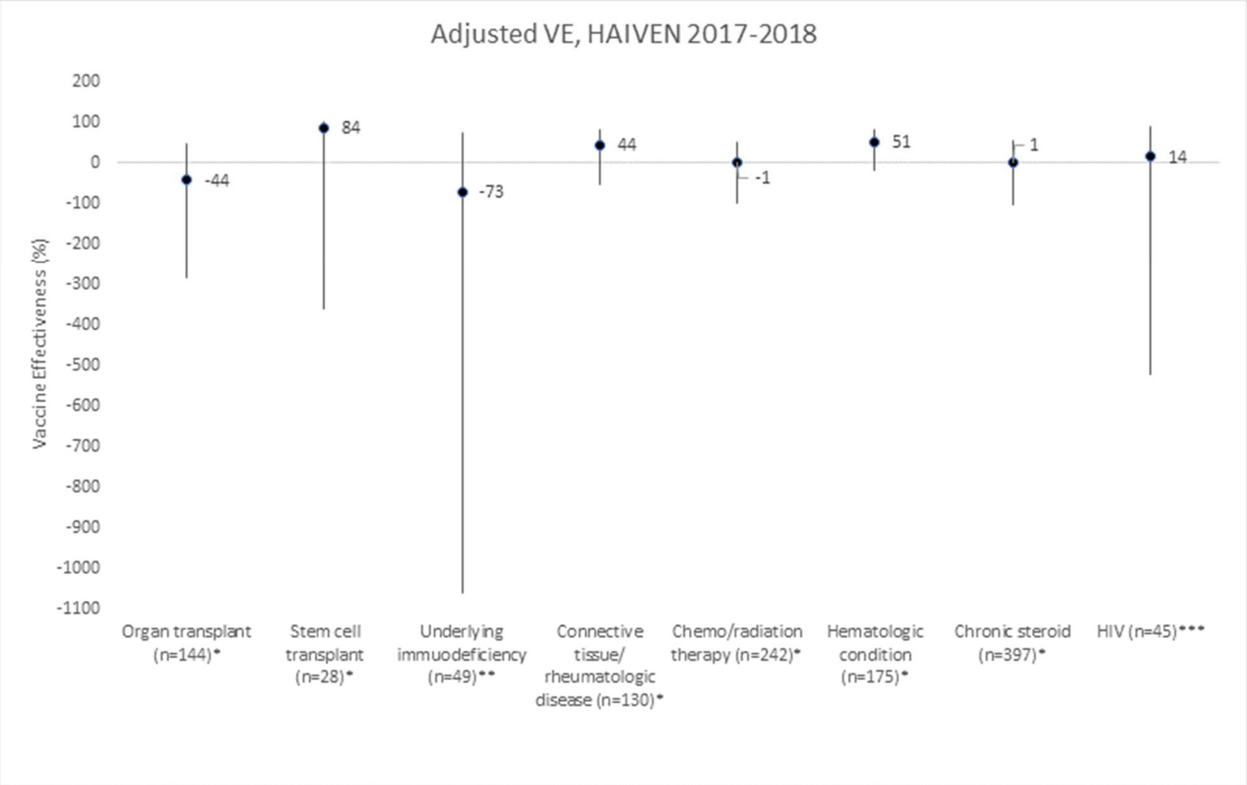

Figure S2 legend:

*Adjusted for enrolling site, onset date (pre-peak, peak, post-peak), age, race, days from illness onset to specimen collection (0-1, 2-4, 5-10 days), self-reported health (poor/fair, good/very good/excellent), self-reported hospitalizations

**Adjusted for enrolling site, onset date (pre-peak, peak, post-peak), age, days from illness onset to specimen collection (0-1, 2-4, 5-10 days), self-reported health (poor/fair, good/very good/excellent), self-reported hospitalizations because the full model did not converge

***Adjusted for enrolling site, age because the full model did not converge
